# Upper-room germicidal ultraviolet light reduced respiratory-related absenteeism among pre-school students: an analysis of newly discovered historical data

**DOI:** 10.1101/2022.08.19.22278959

**Authors:** Christopher W. Ryan

## Abstract

**Objectives:** To assess the effect of upper-room germicidal ultraviolet light systems (GUV) on child absenteeism from a preschool.

**Methods:** I fit a generalized linear model for interventional time series to a newly-discovered historical dataset of attendance records from a preschool from 1941 to 1949, where GUV was installed in December 1945.

**Results:** In all but peak summer months, the presence of GUV was associated with a sizable reduction in child absenteeism due to respiratory illnesses of any cause. Odds ratios for the effect ranged from 0.49 to 0.77, depending on season. In all but high summer, model-predicted absenteeism rates with GUV were a third to a half what they were without it.

**Conclusions:** Installation of upper-room germicidal light in a preschool in December 1945 was associated with a signficant and operationally meaningful reduction in absenteeism due to respiratory illness of any cause. While today’s overall epidemiological environment may be different from that of the 1940s, COVID-19 has shown us that the human species remains vulnerable to pandemics of novel respiratory pathogens, to say nothing of annual influenza outbreaks. Thus air disinfection in congregate indoor settings remains a critical issue. Wider use of upper-room germicidal UV systems in schools and preschools may be worthwhile to reduce absenteeism due to illness, and the educational, social, and economic consequences that ensue.

## 1 Introduction

The COVID-19 pandemic has brought renewed urgency to the problem of indoor air disinfection, particularly in schools. Schools around the world have struggled with proactive closures intended to slow the spread of the pandemic, and with absenteeism among students and staff due to isolation and quarantine.

Consistent school attendance is almost universally considered important for children’s educational, emotional, and social development. Schools are woven into the socioeconomic fabric of society, often serving as social supports for students and families. The adverse effects of missing school fall most heavily upon vulnerable families, exacerbating inequities.^1–3^ Large-scale absenteeism interferes with parental work activities that generate household income.^4,5^ The difficulty in operating hospitals during a pandemic when so many of the staff depend upon schools for out-of-home child care has been a poignant example.^6^

Most schools in the United States (US) closed to in-person instruction in spring of 2020 in an attempt to mitigate the pandemic. Since reopening in fall of 2020, COVID-19 mitigation in US schools has emphasized masking, frequent testing, and eventually vaccination. These interventions require substantial compliance with individual behavioral mandates and are not universally welcomed by parents or staff.

According to Frieden’s “health impact pyramid” model, expecting individual behavior change generally yields the least health impact while demanding the most effort.^7^ Population-level interventions would be more efficient. Continuous disinfection of room air is an example.

Ultraviolet-C (UV-C) radiation kills or inactivates a wide variety of micro-organisms in the air, including many pathogenic viruses and bacteria, as demonstrated in both benchtop^8,9^ and room-sized^10–12^ chambers. Fixtures that emit UV-C at a wavelength of 254 nm were developed in the 1930s. Capitalizing on the vertical convection currents present in occupied rooms, properly-designed 254 nm UV fixtures that narrowly collimate their output can be installed high on walls where they disinfect large volumes of room air as it circulates up and down. (These systems will hereafter be referred to as upper-room GUV or simply GUV.) As the UV light is limited to the upper reaches of the room, the occupants are not exposed to any meaningful degree^13–15^ but still benefit from the disinfection of the air they breathe and share with other occupants. GUV has been demonstrated to produce the equivalent of 7 to 150 additional air change equivalents per hour (ACHeq).^16^ This is substantially higher than could practicably be achieved by mechanical ventilation/filtration.

The cyclical history of GUV has been reviewed by Reed.^17^ It was installed in public spaces for a time in the 1940s to early 1950s but then largely disappeared from use in the US. GUV experienced a brief resurgence in US medical facilities in the late 1980s due to nosocomial outbreaks of drug-resistant TB associated with HIV infection. It is rarely found in public spaces in the US today.

In early studies of GUV in schools, the outcome of interest was usually the incidence rates of specific respiratory diagnoses such as measles, chickenpox, or mumps.^18–20^ The experimental designs differed, and the results were mixed. These studies were limited by the analytical methods available at the time.

Notwithstanding specific diagnoses, attendance (or its converse, absenteeism), is of great concern to students, families, and school administrators. There is little published evidence concerning the practical effect of upper-room GUV on school attendance. In 1950, Gelperin and colleagues in New Haven, Connecticut installed GUV in some classrooms in each of eight out of 36 buildings in February. They concluded there was no discernible effect on absenteeism rates due to respiratory illnesses, but the observation period was only 4.5 months–less than one school year.^21^ More recently, Su, et al found no difference in absenteeism rates, in a single elementary school, between students in two classrooms with GUV compared to those in four classrooms without. However, their primary interest was the comparative efficiency of two different air sampling techniques; absenteeism rates were peripheral to their study objectives. Again, the observation periods were short. The data were sparse, the non-parametric methods used for analysis had little power, and serial autocorrelation between months was not accounted for.^22^

A prospective randomized trial using modern methods would be ideal, but conducting such a study in school buildings would face many design, analysis, logistical, and political hurdles. The serendipitous discovery of several historical documents in the archives of a major current-day manufacturer of GUV systems presents an opportunity to assess the practical effect of upper-room GUV on student absenteeism in a preschool over a span of nine years, using modern analytical methods.

## 2 Methods

### 2.1 The source document and data

The documents were discovered in the archives of Atlantic Ultraviolet. Most seemed to be internal documents generated by, or under the auspices of, GUV manufacturers or vendors, in particular Westinghouse (makers of a GUV lamp called “Sterilamp”) and a company called Sanitron, a predecessor of Atlantic Ultraviolet. Atlantic Ultraviolet was founded in the early 1960s and was the sole east coast distributor for Westinghouse’s Sterlamps. In the late 1960s they distanced themselves somewhat from Westinghouse and began manufacturing their own ultraviolet units, for both air and water purification (personal communication, Ann Wysocki, Director of Marketing, Atlantic Ultraviolet Corporation). Most of the documents are anonymous. Of those that display a date, they range from 1946 to 1950. One document mentions that it was published in the New York State Journal of Medicine and presented at the 140th annual meeting of the Medical Society of the State of New York, section on pediatrics, on May 2, 1946; a Medline search confirmed that.^23^

The document of most interest for the present study is entitled “Reduction in absentee rate at the [preschool A] using Westinghouse germicidal Sterilamps.” Preschool A is located in what was at the time a moderately-large Mid-Atlantic US city. The document displays no author, affiliation, or other clues as to its originator. Although Preschool A is still in operation, I know little about how it functioned 70 years ago. The document contains only one detail: that in the summer months, the children spent a lot of time outdoors.

The document includes a table of attendance data from January 1941 through November 1949, inclusive. Recorded are the number of possible/available child-days of attendance and the number of child-days missed due to respiratory illness, for each age group each month. There are four age groups: ages 2, 3, 4, and 5. The five-year-olds were considered to be in a kindergarten program that did not operate in July or August. For the younger age groups, attendance figures are recorded year-round, except for four summers (July and August), in which there was “no group” for four-year-olds. “Sterilamp” GUV systems were installed in all classrooms in December 1945. The document does not explain why, but other historical documents indicate Preschool A often participated in research studies. Analytically, the document contains just one simple graph (reproduced in the Supplemental Materials) highly suggestive of a reduction of absenteeism associated with the presence of GUV. There was no apparent attempt to fit a statistical model, account for serial autocorrelation in this time series, conduct hypothesis tests, or construct confidence intervals.

Using the detailed table of attendance data, I fit an interventional times series model in an effort to measure the effect of the upper-room GUV systems on student absenteeism.

### 2.2 Statistical modeling strategy

Without knowing how much the different age groups interacted, I pooled the data across age groups and analyzed absenteeism in a school-wide fashion. I calculated monthly absenteeism rates by dividing the number of child-days missed by the number of child-days available for each month, yielding a proportion of available days missed in each month. I used the logit (log-odds) transformation of those absenteeism rates as the response. I assumed an autoregressive lag-1 correlation structure between the monthly observations. Based on exploratory graphs, I allowed for two different homoskedastic variance structures, one before GUV was installed and one after. The predictor of interest was the presence/absence of upper-room GUV. I assumed the fixtures were installed at the end of December 1945 and counted January 1946 as the first month with GUV.

The effect of GUV, if any, on preschool absenteeism due to respiratory illnesses could reasonably be expected to vary seasonally. The baseline incidence of such illnesses is higher in colder months, so I used each month’s average temperature as a proxy for those seasonal fluctuations. It also served to represent seasonal cycles in where children might have spent their time while in school, indoors versus out. I obtained temperature data from January 1941 through November 1949, inclusive, from the National Weather Service here: https://www.weather.gov/wrh/Climate?wfo=lwx on 8 July 2022.

Confounding between the passage of time and the effect of an intervention instituted at a specific point in time is a challenging aspect of interventional times series analyses. Effects on the response that appear to be associated with the intervention may, at least in part, be due to secular trends or unmeasured temporal factors. To determine whether there were temporal trends in absenteeism unrelated to any putative effects of GUV, I initially included as predictors two piecewise-linear temporal trends, one before GUV was installed and one after, with time measured as the number of months elapsed since the start of the time series in January 1941. I tested the significance of these temporal trend terms.

After fitting the model, I explored customary regression diagnostic plots. I tested hypotheses concerning the effect of GUV on absenteeism, and its interaction with temperature as a proxy for seasonal variations in respiratory epidemiology and child outdoor activity.

I conducted the analysis in R.^24^ My R code, and a pipe-delimited plain-text file containing the complete data, are both available in the Supplemental Materials.

### 2.3 Reviews and approvals

The SUNY Upstate Medical University Institutional Review Board determined that this project is not human subject research because it relies only on aggregated, non-identifiable data.

## 3 Results

### 3.1 Data exploration

After pooling across age groups, the analytical dataset comprised 107 monthly observations. There were 59 months without upper-room GUV, followed by 48 with GUV. Monthly respiratory-related absenteeism rates ranged from 1.7% to 37.5%, with a median of 8.7%. Monthly average temperatures ranged from 28.8 F to 81 F, with a median of 57.7 F. Temperatures varied seasonally as expected.

As shown in Figures 1 and 2, extremes of absenteeism rates seem to be more marked in winter months, and these extremes may have been blunted during the GUV years, resulting in less variation in absenteeism rates during the GUV period.

**Figure 1:**
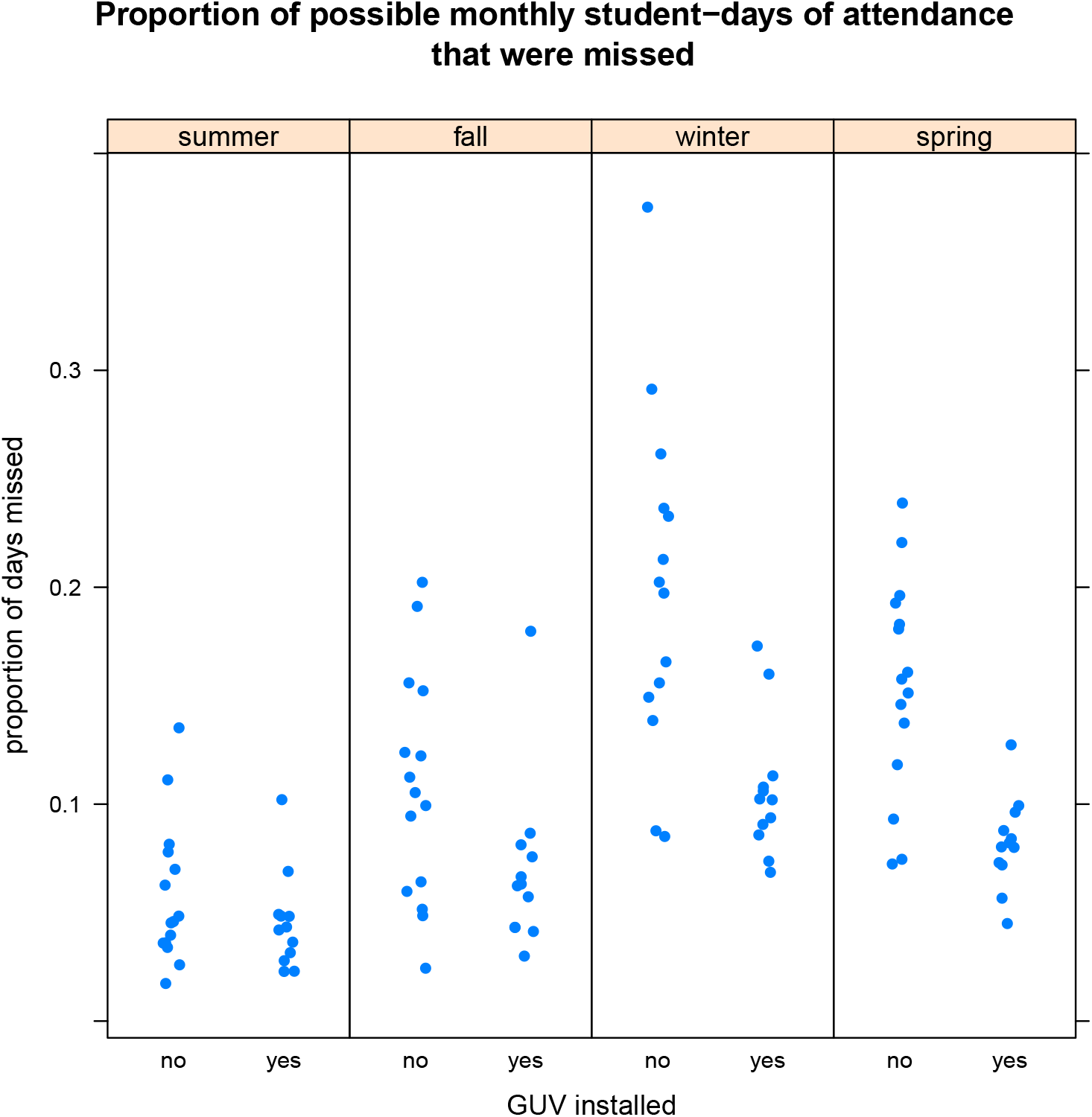
Monthly absenteeism rates, grouped by season. Not surprisingly, extremes of absenteeism seemed to occur more commonly in fall, winter, and spring

**Figure 2:**
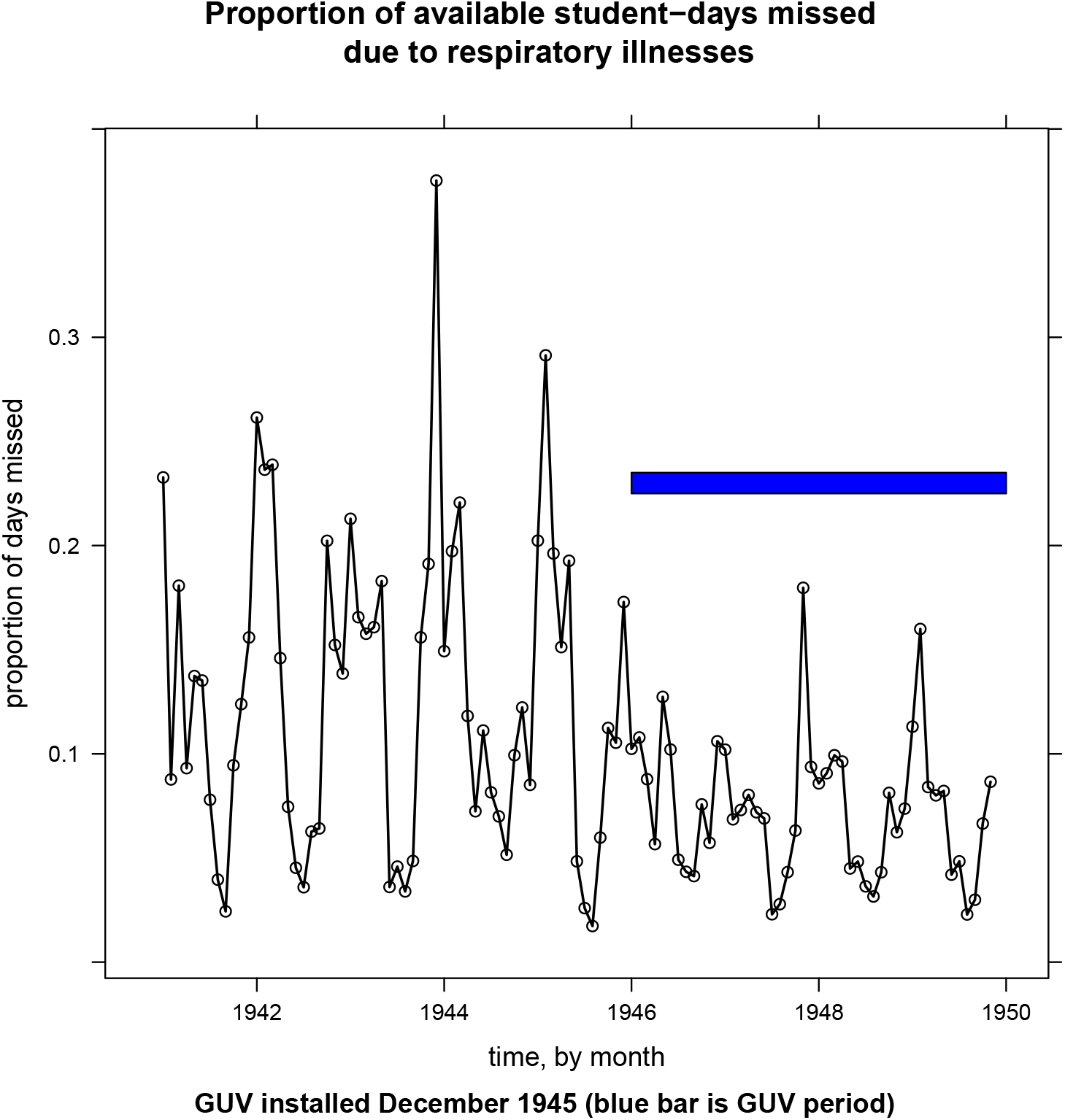
Monthly absenteeism rate for all age groups combined. The blue bar indicates the GUV period. The distribution of absenteeism rates seems compressed to a lower ranged during the GUV period.

**Figure 3:**
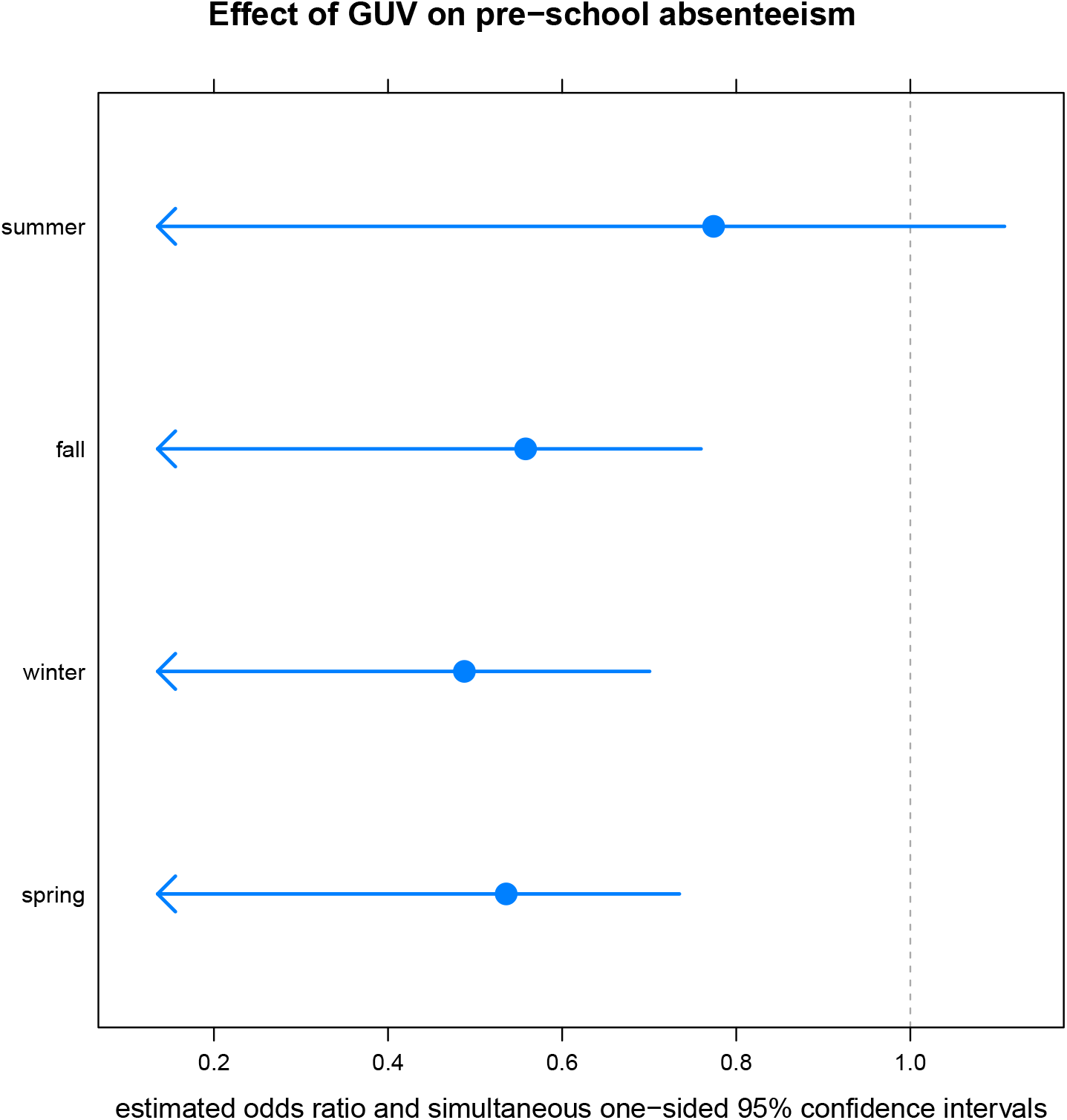
Odds ratios for reduction in any-respiratory-cause absenteeism from a preschool by upper-room germicidal light, stratified by the mean temperature of each of the four seasons

### 3.2 Modeling

Initial modeling indicated that the effect of temperature on absenteeism might be quadratic, so I also included the square of average monthly temperature as a predictor, along with its interaction with GUV. Details of several models and residual diagnostic plots are shown in the Supplemental Materials.

Whether any time trends were needed in the model was a critical question. In Model (1), the coefficient on the time index term represents the linear effect of time before the installation of GUV. The sum of that coefficient plus the coefficient on the guv:time interaction term represents the linear effect of time after the installation of GUV. A two-sided test failed to reject, at the 0.05 level, the null hypothesis that these two time trends were simultaneously zero (p-values 0.54 and 0.47 for the pre-GUV and post-GUV time trends, respectively.) Qualitatively, this comports with Figure 2. Therefore I chose to remove all time-related terms from the mean structure of the model, but I retained the AR(1) autocorrelation. Thus Model (4) in the Supplementary Materials is the “working model” for all analyses, diagnostics, and interpretation henceforth. Model (4) fits best by AIC but is not much different in that regard from Models (1), (3), and (6).

Diagnostic plots for the working model were generally reassuring, except for some remaining autocorrelation and seasonality, especially at a lag of around 4 months.

### 3.3 Interpretation of the working model

Odds ratios, and their simultaneous one-sided 95% confidence intervals, for the reduction of absenteeism by GUV at the mean temperature of each conventional 3-month season are shown in Table 3. At non-summer temperatures, the ORs are all approximately 0.5 to 0.6 and suggest a significant reduction in any-cause respiratory absenteeism.

Figure 4 shows the reduction in respiratory-related absenteeism in the presence of GUV, and how that effect varies with monthly average temperature (as a surrogate representing seasonal variations in respiratory epidemiology and in child activities.) Save for peak summer temperatures, absenteeism rates due to any-cause respiratory illness are significantly and meaningfully lower when upper-room GUV is present.

**Figure 4:**
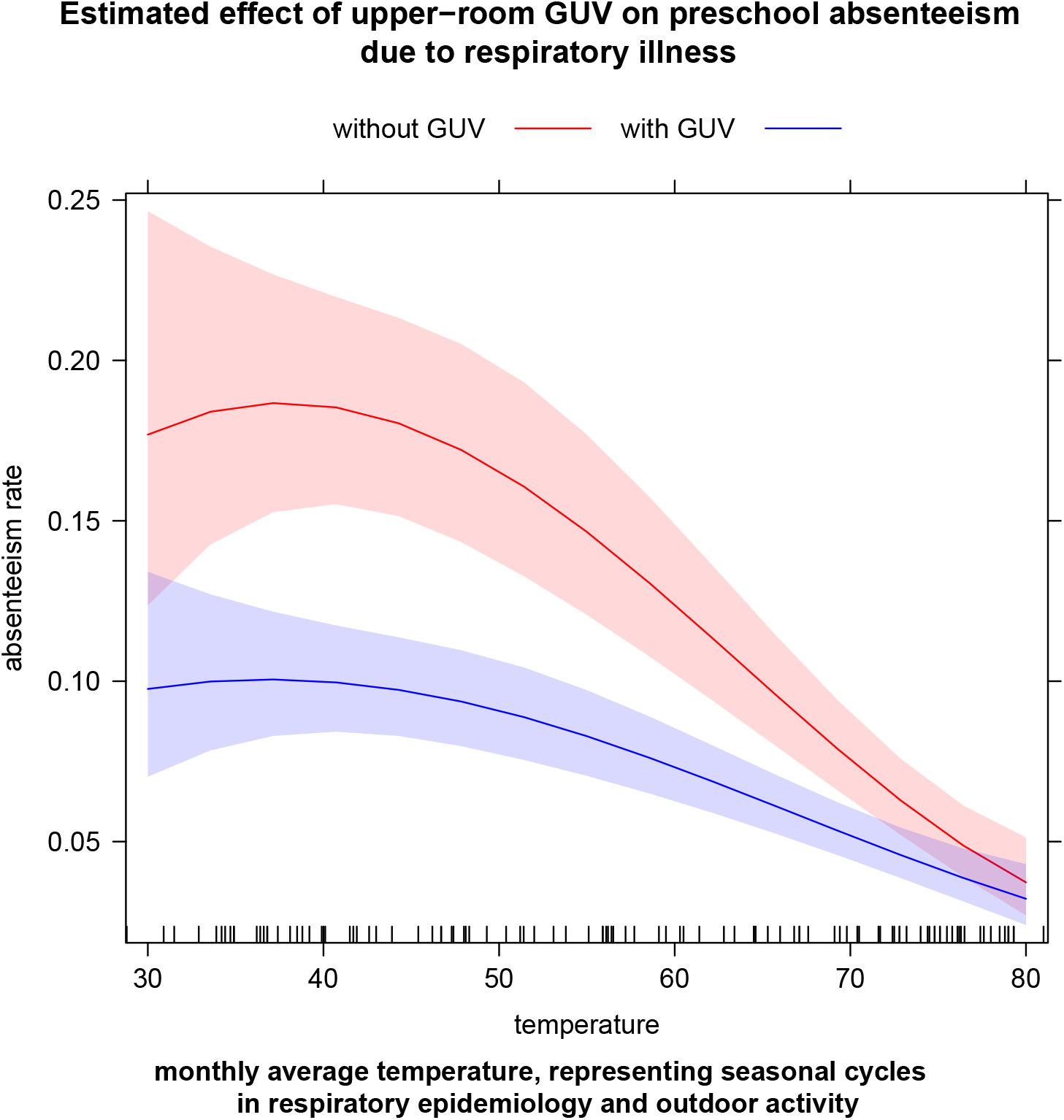
Solid lines show model-estimated monthly child absenteeism rates from pre-school due to respiratory illness before and after upper-room GUV was installed. Shaded regions are 95% pointwise confidence bands for each estimate. Red represents the months prior to GUV, while blue represents months with GUV. The effect varies by monthly average temperature (used as a proxy for seasonal cycles in respiratory epidemiology and in child outdoor activity), but GUV reduces predicted absenteeism by nearly half at all but the highest summer temperatures.

Figure 5 illustrates the observed and model-predicted monthly absenteeism rate, and the counterfactual: the model-predicted upper respiratory absenteeism rate had upper room GUV *not* been installed in December 1945.

**Figure 5:**
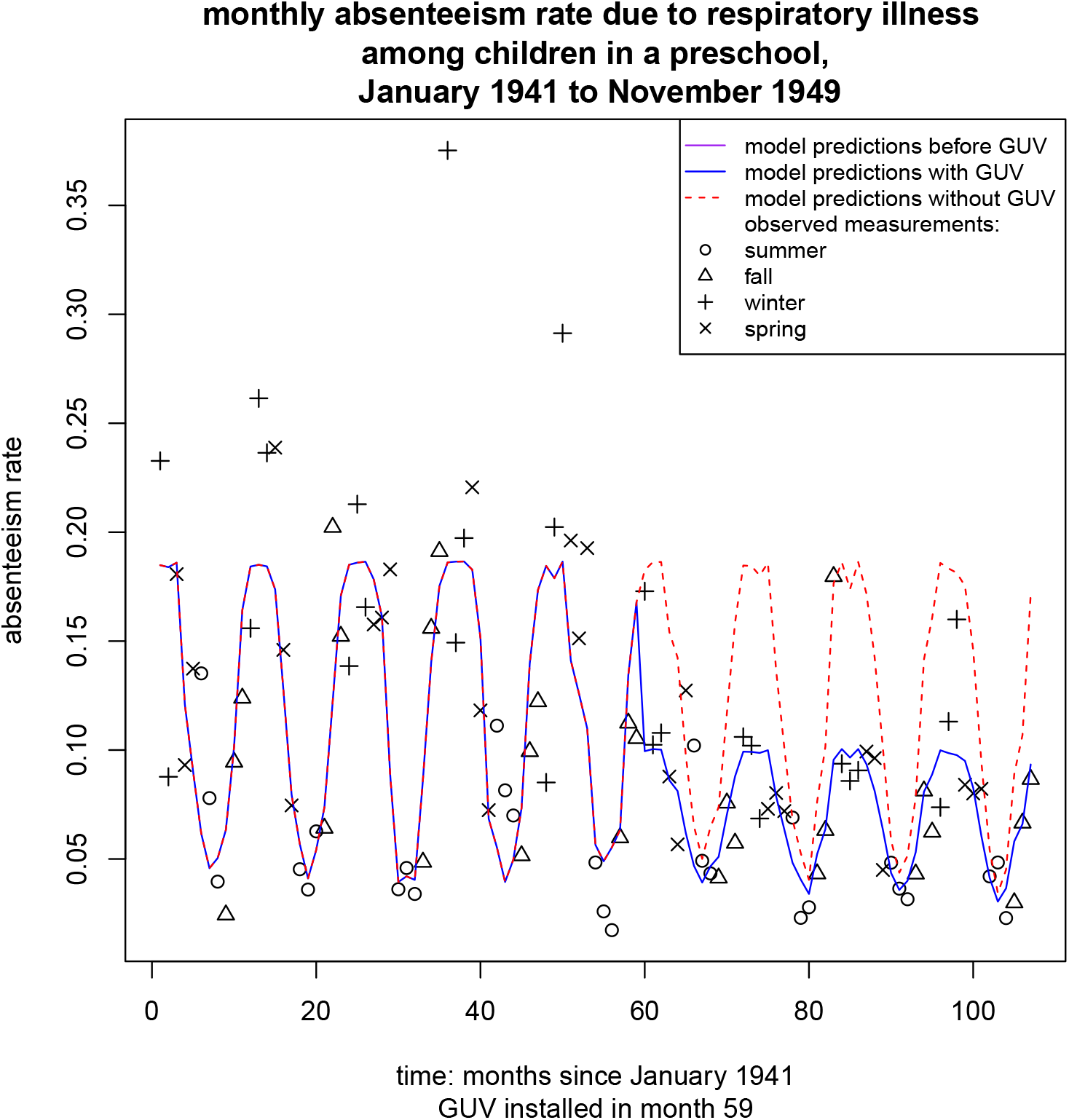
Time series of observed and model-predicted monthly absenteeism rates, and the model-predicted rates had GUV *not* been installed in December 1945 (dotted red line). The first complete month with GUV was January 1946 (month 60).

## 4 Discussion

This analysis demonstrated a sizable reduction in child absenteeism from preschool due to respiratory illnesses of any cause, associated with the installation of upper-room 245 nm germicidal ultraviolet lights. In all but peak summer months, absenteeism rates with GUV were a third to a half of what they were without it. The reduction appeared to be abrupt, concurrent with the installation of GUV systems in December 1945. There was no discernible downward temporal trend in absenteeism either before or after the installation.

Irrespective of specific diagnosis, all-cause absenteeism from preschool is of great concern to families, school administrators, and employers. Respiratory syndromes are one of the most common causes of absenteeism. Thus a reduction in respiratory-related school absences would be important to all parties. To my knowledge, this analysis of nine years of comprehensive attendance data from a preschool is the first to demonstrate such a reduction.

Previous studies of upper-room GUV in schools focused on the incidence of specific respiratory diagnoses. Effects on overall incidence were not always convincing. In some of those studies, there may however have been a “blunted” epidemiologic pattern in settings with GUV: a lower incidence rate drawn out over a longer period of time.^20^ Whether this would be valued by schools or families is unknown.

Designs in which all classrooms in a building were outfitted with GUV and a separate building used as a control seemed to yield more encouraging results in reducing disease incidence than those in which irradiated classrooms and control classrooms were present in the same building. Examples of this design include Swarthmore public elementary school versus high school under Wells^18^ and Cato-Meridian public schools versus Mexico public schools under Perkins and Bahlke^19,20^

The studies by Gelperin^21^ and Su^22^ represent the alternative design with”internal controls,” meaning some rooms in a school building were outfitted with GUV while others in the same building were not. Over short observation periods, neither reported a difference in absenteeism associated with GUV. Given the inevitable interpersonal interactions between children and teachers throughout a school, it is worth considering whether the measurable effect of GUV may depend on whether all classrooms in a building are treated. The analysis presented here was of a whole-building GUV installation with a much longer observation period than any previous published work, and showed a meaningful reduction in absenteeism.

### 4.1 Limitations

Interventional time series analyses are subject to a number of limitations. In the absence of a concurrent control group, it is impossible to say with certainty that the putative effect of the intervention was not due to some other factor or event that occurred around the same time. The occurrence of such a factor or event is particularly difficult to discern at a 70-year remove.

Furthermore, the overall community incidence of respiratory illnesses in children can vary cyclically across years. It is possible that the non-GUV years early in this time series were unusually “bad” in that regard, and that the decrease in the subsequent GUV years was just part of that natural cycle. It is even possible that the administrators at Preschool A decided to install GUV in December 1945 *because* they had just experienced several years of high levels of absenteeism. It is always important to remember that any time series is only a segment of a longer series that precedes and follows it. However, I found no historical references to particularly severe outbreaks of respiratory diseases in the pre-GUV years of 1941 to 1945. In fact, a notable influenza epidemic was reported in the 1946-1947 season.^25–27^ Yet the absenteeism rate during that 1946-1947 influenza season, during which Preschool A had GUV, was significantly lower than in the preceding five years.

No information is available in the source document about how absenteeism due to respiratory illness was defined and counted, nor by whom. Likewise, there is no information about adverse effects attributable to the GUV.

## 5 Conclusions

It is difficult to extrapolate from the overall epidemiologic environment of the 1940s to that of the present. However, as the COVID-19 pandemic demonstrates, the human species remains vulnerable to pandemics of novel respiratory pathogens, to say nothing of the well-known annual influenza season. Thus air disinfection in congregate indoor settings remains a critical issue. Wider use of upper-room germicidal UV systems in schools and preschools may be worthwhile to reduce absenteeism due to illness, and the educational, social, and economic consequences that ensue.

## Supporting information

R code for analysis (extension can be changed to .R)

Pipe-delimited plain-text file with the data

Additional graphs and model diagnostic plots

## Data Availability

All data used in this analysis are included in the Supplemental Materials as a pipe-delimited plain-text file.

## 6 Acknowledgments

I wish to thank Chloe Parker of the Broome County Health Department for data entry; Dr. Mei-Hsiu Chen of the Binghamton University Statistical Consulting Service for helpful suggestions on tatistical modeling; and Heather Ryan of the Broome County Health Department for historical research and editorial assistance with the manuscript.

## 7 Declaration of conflicting interests

Competing interests: All authors have completed the ICMJE uniform disclosure form at www.icmje.org/coi_disclosure.pdf and declare: no support from any organization for the submitted work; no financial relationships with any organizations that might have an interest in the submitted work in the previous three years; no other relationships or activities that could appear to have influenced the submitted work.

## Notes

### Competing Interest Statement

The authors have declared no competing interest.

### Funding Statement

This study did not receive any funding.

### Author Declarations

The SUNY Upstate Medical University Institutional Review Board determined that this project is not human subject research because it relies only on aggregated, non-identiable data.

## References

[1] UNESCO. Education: From disruption to recovery. United Nations Educational, Scientific, and Cultural Organization. June 29, 2022. URL: https://en.unesco.org/covid19/educationresponse#schoolclosures.

[2] R. Viner et al. “School Closures During Social Lockdown and Mental Health, Health Behaviors, and Well-being Among Children and Adolescents During the First COVID-19 Wave: A Systematic Review.” In: JAMA pediatrics 176 (4 Apr. 2022), pp. 400–409. ISSN: 2168-6211. doi: 10.1001/jamapediatrics.2021.5840. ppublish.

[3] R. Lordan et al. “Considerations for the Safe Operation of Schools During the Coronavirus Pandemic.” In: Frontiers in public health 9 (2021), p. 751451. ISSN: 2296-2565. doi: 10.3389/fpubh.2021.751451. epublish.

[4] R. M. Viner et al. “School closure and management practices during coronavirus outbreaks including COVID-19: a rapid systematic review.” In: The Lancet. Child and adolescent health 4 (5 May 2020), pp. 397–404. ISSN: 2352-4650. doi: 10.1016/S2352-4642(20)30095-X. ppublish.

[5] M. R. Keogh-Brown et al. “The macroeconomic impact of pandemic influenza: estimates from models of the United Kingdom, France, Belgium and The Netherlands.” In: The European journal of health economics : HEPAC : health economics in prevention and care 11 (6 Dec. 2010), pp. 543–554. ISSN: 1618-7601. doi: 10.1007/s10198-009-0210-1. ppublish.

[6] J. Bayham and E. P. Fenichel. “Impact of school closures for COVID-19 on the US health-care workforce and net mortality: a modelling study.” In: The Lancet. Public health 5 (5 May 2020), e271–e278. ISSN: 2468-2667. doi: 10.1016/S2468-2667(20)30082-7. ppublish.

[7] T. R. Frieden. “A framework for public health action: the health impact pyramid.” eng. In: Am J Public Health 100.4 (2010), pp. 590–595. doi: 10.2105/AJPH.2009.185652. URL: http://dx.doi.org/10.2105/AJPH.2009.185652.

[8] C. M. Walker and G. Ko. “Effect of ultraviolet germicidal irradiation on viral aerosols.” In: Environmental science & technology 41 (15 Aug. 2007), pp. 5460–5465. ISSN: 0013-936X. doi: 10.1021/es070056u.

[9] D.-K. Kim and D.-H. Kang. “UVC LED Irradiation Effectively Inactivates Aerosolized Viruses, Bacteria, and Fungi in a Chamber-Type Air Disinfection System.” In: Applied and environmental microbiology 84 (17 Sept. 2018). ISSN: 1098-5336. doi: 10.1128/AEM.00944-18.

[10] M. First et al. “Fundamental factors affecting upper-room ultraviolet germicidal irradiation - part I. Experimental.” In: Journal of occupational and environmental hygiene 4 (5 May 2007), pp. 321–331. ISSN: 1545-9624. doi: 10.1080/15459620701271693.

[11] C. Su, J. Lau, and F. Yu. “A Case Study of Upper-Room UVGI in Densely-Occupied Elementary Classrooms by Real-Time Fluorescent Bioaerosol Measurements”. In: International Journal of Environmental Research and Public Health 14.1 (Jan. 2017), p. 51. ISSN: 1660-4601. doi: 10.3390/ijerph14010051. URL: http://www.mdpi.com/1660-4601/14/1/51.

[12] S. L. Miller et al. Efficacy of ultraviolet irradiation in controlling the spread of tuberculosis. Research rep. University of Colorado Boulder, Oct. 19, 2002.

[13] E. A. Nardell et al. “Safety of upper-room ultraviolet germicidal air disinfection for room occupants: results from the Tuberculosis Ultraviolet Shelter Study.” In: Public health reports (Washington, D.C. : 1974) 123 (1 2008), pp. 52–60. ISSN: 0033-3549. doi: 10.1177/003335490812300108. ppublish.

[14] M. W. First et al. “Monitoring human exposures to upper-room germicidal ultraviolet irradiation.” In: Journal of occupational and environmental hygiene 2 (5 May 2005), pp. 285–292. ISSN: 1545-9624. doi: 10.1080/15459620590952224. ppublish.

[15] UV-C Photocarcinogenesis Risks From Germicidal Lamps. Tech. rep. CIE 187:2010. International Commission on Illumination, 2010. isbn: ISBN 978 3 901906 81 7.

[16] J. J. McDevitt et al. “Inactivation of poxviruses by upper-room UVC light in a simulated hospital room environment.” In: PloS one 3 (9 Sept. 2008), e3186. ISSN: 1932-6203. doi: 10.1371/journal.pone.0003186. epublish.

[17] N. G. Reed. “The history of ultraviolet germicidal irradiation for air disinfection.” In: Public health reports (Washington, D.C. : 1974) 125 (1 2010), pp. 15–27. ISSN: 0033-3549. doi: 10.1177/003335491012500105.

[18] W. F. Wells, M. W. Wells, T. S. Wilder, et al. “The Environmental Control of Epidemic Contagion. I. An Epidemiologic Study of Radiant Disinfection of Air in Day Schools.” In: American Journal of Hygiene 35.1 (1941), pp. 97–121.

[19] J. E. Perkins, A. M. Bahlke, and H. F. Silverman. “Effect of ultra-violet irradiation of classrooms on spread of measles in large rural central schools.” In: American journal of public health and the nation’s health 37 (5 May 1947), pp. 529–537. ISSN: 0002-9572.

[20] A. M. Bahlke, H. F. Silverman, and H. S. Ingraham. “Effect of ultra-violet irradiation of classrooms on spread of mumps and chickenpox in large rural central schools.” In: American journal of public health and the nation’s health 39 (10 Oct. 1949), pp. 1321–1330. ISSN: 0002-9572. doi: 10.2105/ajph.39.10.1321. ppublish.

[21] A. Gelperin, M. A. Granoff, and J. I. Linde. “The effect of ultraviolet light upon absenteeism from upper respiratory infections in New Haven schools.” In: American journal of public health and the nation’s health 41 (7 July 1951), pp. 796–805. ISSN: 0002-9572. doi: 10.2105/ajph.41.7.796. ppublish.

[22] C. Su, J. Lau, and S. Gibbs. “Student absenteeism and thecomparisons of two samplingprocedures for culturable bioaerosolmeasurement in classrooms with andwithout upper room ultravioletgermicidal irradiation devices”. In: Indoor and Built Environment 25.3 (2016), pp. 551–562. doi: 10.1177/1420326X14562257. URL: https://journals.sagepub.com/doi/10.1177/1420326X14562257.

[23] R. A. Higgons and H. Y. D. E. G M. “Effect of ultraviolet air sterilization upon incidence of respiratory infections in a children’s institution; a 6-year study.” In: New York state journal of medicine 47 (7 Apr. 1947), pp. 707–710. ISSN: 0028-7628. ppublish.

[24] R Core Team. R: A Language and Environment for Statistical Computing. R Foundation for Statistical Computing. Vienna, Austria, 2022. URL: https://www.R-project.org/.

[25] J. Salk and P. Surianao. “Importance of antigenic composition of influenza virus vaccine in protecting against the natural disease; observations during the winter of 1947-1948.” In: American journal of public health and the nation’s health 39 (3 Mar. 1949), pp. 345–355. ISSN: 0002-9572. doi: 10.2105/ajph.39.3.345. ppublish.

[26] K. ED. and L. O. G. E. J P. “Influenza A prime: a clinical study of an epidemic caused by a new strain of virus.” In: Annals of internal medicine 33 (2 Aug. 1950), pp. 371–379. ISSN: 0003-4819. doi: 10.7326/0003-4819-33-2-371. ppublish.

[27] E. D. Kilbourne et al. “The total influenza vaccine failure of 1947 revisited: major intrasubtypic antigenic change can explain failure of vaccine in a post-World War II epidemic.” In: Proceedings of the National Academy of Sciences of the United States of America 99 (16 Aug. 2002), pp. 10748–10752. ISSN: 0027-8424. doi: 10.1073/pnas.162366899. ppublish.

