## Additional graphs and model diagnostic plots for "Upper-room germicidal ultraviolet light reduced respiratory-related absenteeism among pre-school students: an analysis of newly discovered historical data"

Christopher W. Ryan, MD, MS, MSPH

August 18, 2022

Figure 1 is the single graph appearing in the original historical document (with the name of the preschool redacted).

Figure 2 shows the expected periodicity in monthly average temperature.

Table 1 compares several models fit to the data. All models were fit via generalized least squares, allowing for different variances under pre-GUV and post-GUV periods, and with auto-regressive (1-month lag) correlation structure within pre- and post-GUV periods. As expected in an interventional time series analysis, the intervention (installation of GUV) is confounded with the passage of time. When GUV and time are both included (Model (1)), there is no appreciable temporal trend in absenteeism during either pre- or post-GUV periods. Therefore, on statistical and substantive grounds, Model (4), without terms for time, is considered the working model; it also fits best by AIC.

Figure 3 shows the need for a quadratic temperature term.

Figures 4 through 7 are diagnostic plots from the working model, Model (4) in Table 1.

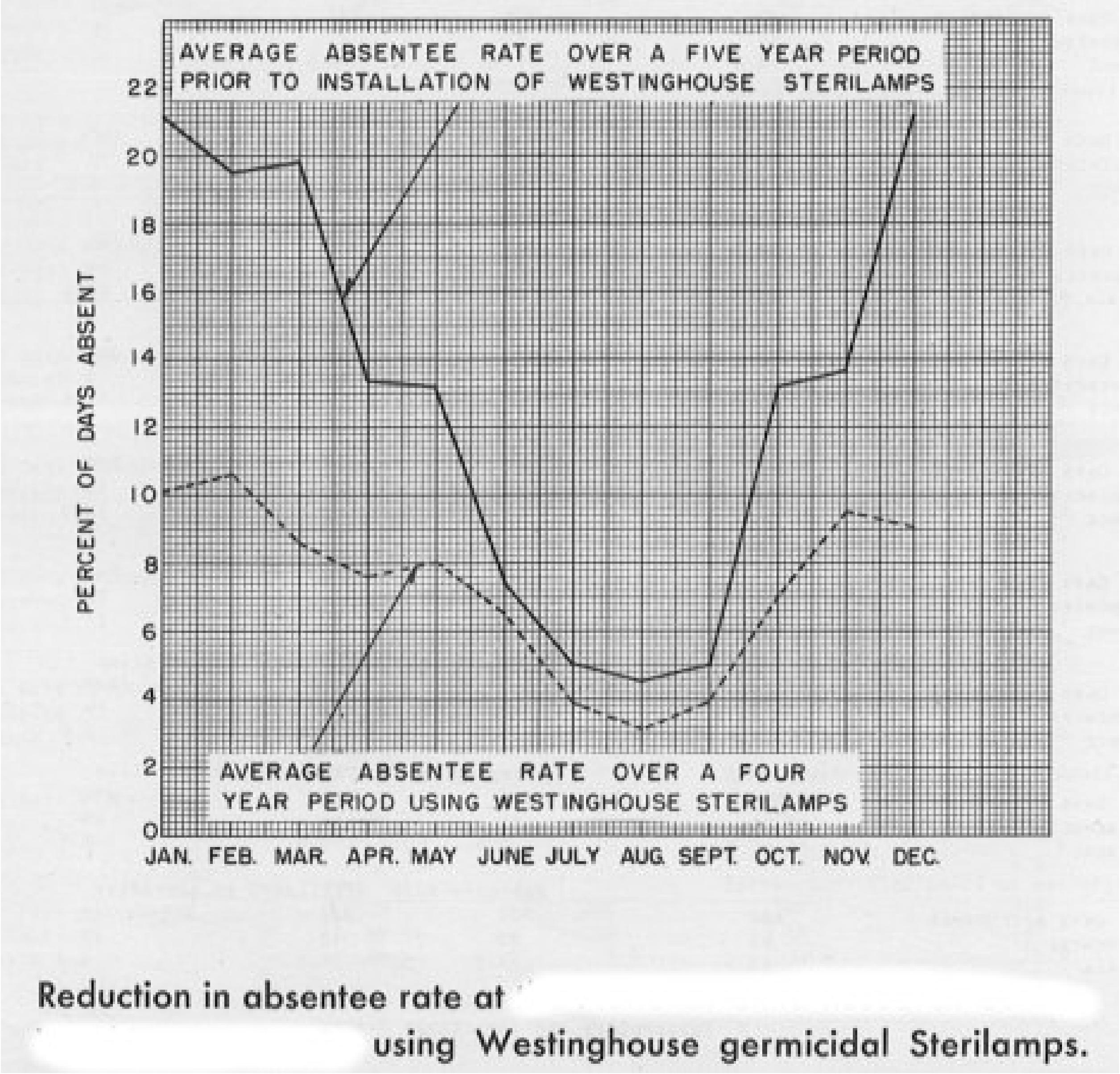


Figure 1: The single graph that appeared in the original document entitled "Reduction in absentee rate at the [pre-school A] using Westinghouse germicidal Sterilamps."

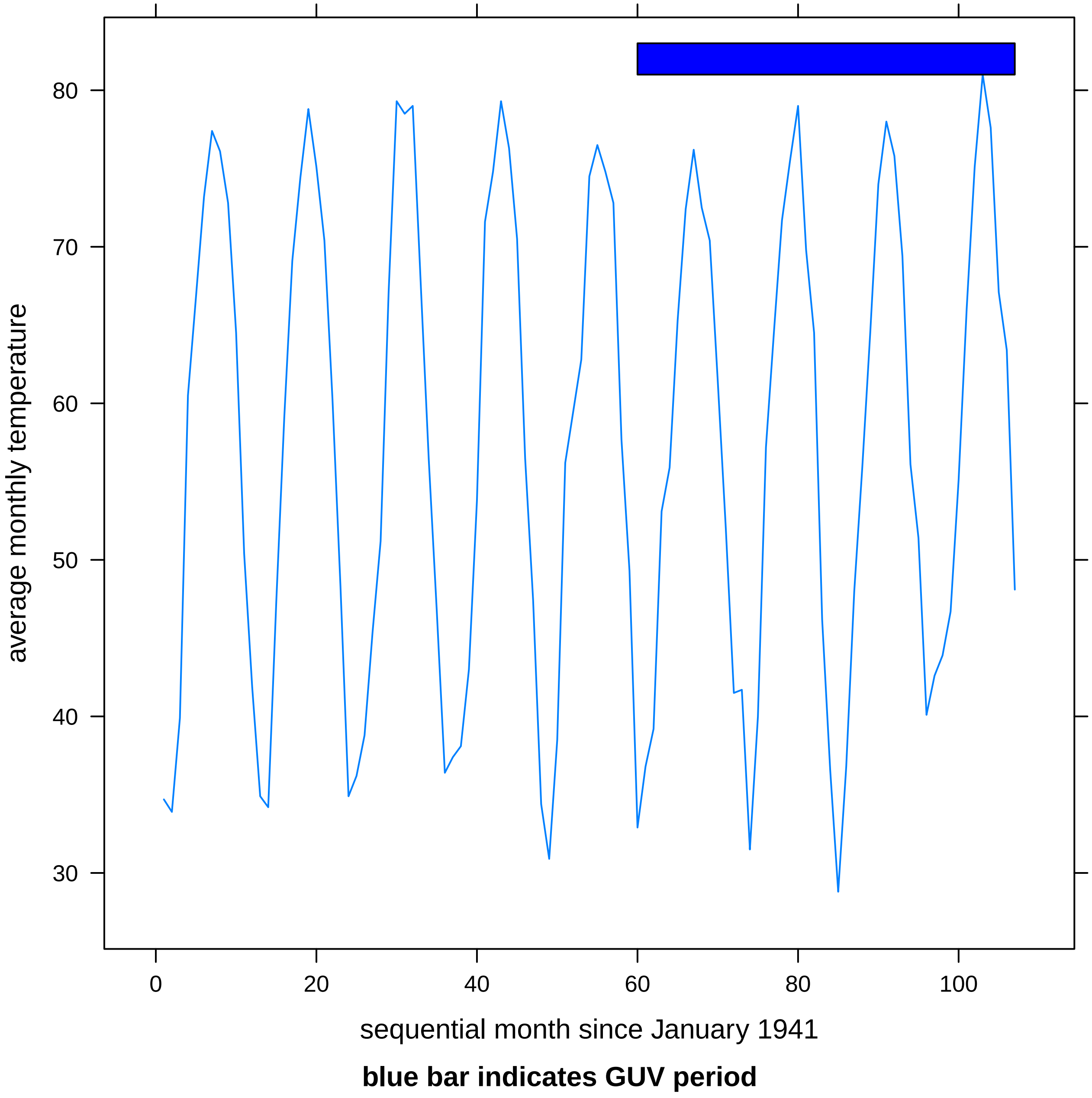


Figure 2: Monthly average temperatures during study period, showing the expected seasonal cycles. Blue bar indicates the period when GUV was present in the classrooms.

Table 1: Model comparison

| | *Dependent variable:* | | | | | |
|---|---|---|---|---|---|---|
| | log-odds of monthly absenteeism rate | | | | | |
| | (1) | (2) | (3) | (4) | (5) | (6) |
| Constant | −2.921 | 0.047 | −2.675 | −2.932 | −0.862 | −2.711 |
| | (1.064) | (0.320) | (0.692) | (1.060) | (0.209) | (0.692) |
| guv.fac1 | 0.163 | −0.889 | | −0.157 | −0.571 | −0.242 |
| | (1.397) | (0.552) | | (1.372) | (0.449) | (0.429) |
| temperature | 0.079 | −0.037 | 0.065 | 0.077 | | 0.063 |
| | (0.040) | (0.005) | (0.026) | (0.040) | | (0.026) |
| time.index | −0.003 | −0.002 | −0.008 | | −0.002 | −0.003 |
| | (0.004) | (0.005) | (0.002) | | (0.005) | (0.005) |
| I(temperatureˆ2) | −0.001 | | −0.001 | −0.001 | −0.0003 | −0.001 |
| | (0.0004) | | (0.0002) | (0.0004) | (0.00004) | (0.0002) |
| guv.fac1:temperature | −0.027 | 0.013 | | −0.028 | | |
| | (0.052) | (0.007) | | (0.051) | | |
| guv.fac1:time.index | −0.003 | −0.003 | | | −0.003 | −0.001 |
| | (0.006) | (0.007) | | | (0.007) | (0.007) |
| guv.fac1:I(temperatureˆ2) | 0.0003 | | | 0.0004 | 0.0001 | |
| | (0.0005) | | | (0.0005) | (0.0001) | |
| Observations | 107 | 107 | 107 | 107 | 107 | 107 |
| Log Likelihood | −51.701 | −58.735 | −55.351 | −52.654 | −55.077 | −54.068 |
| Akaike Inf. Crit. | 125.402 | 135.469 | 124.703 | 123.308 | 128.154 | 126.135 |
| Bayesian Inf. Crit. | 154.803 | 159.525 | 143.413 | 147.363 | 152.209 | 150.191 |

*Note:*

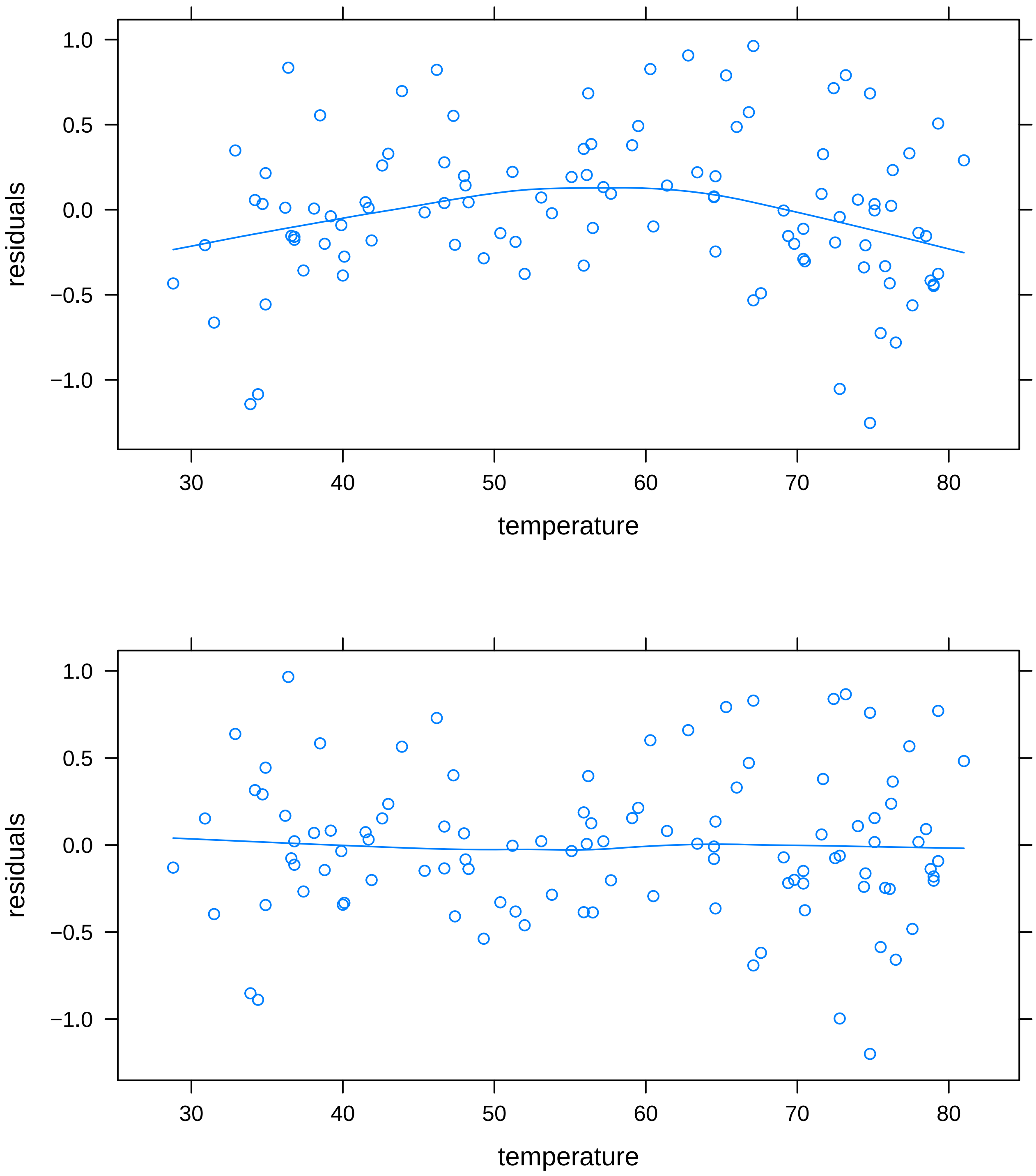


Figure 3: Inclusion of a quadratic term (bottom) eliminates a pattern otherwise found in the residuals from the working model with respect to monthly average temperature (top).

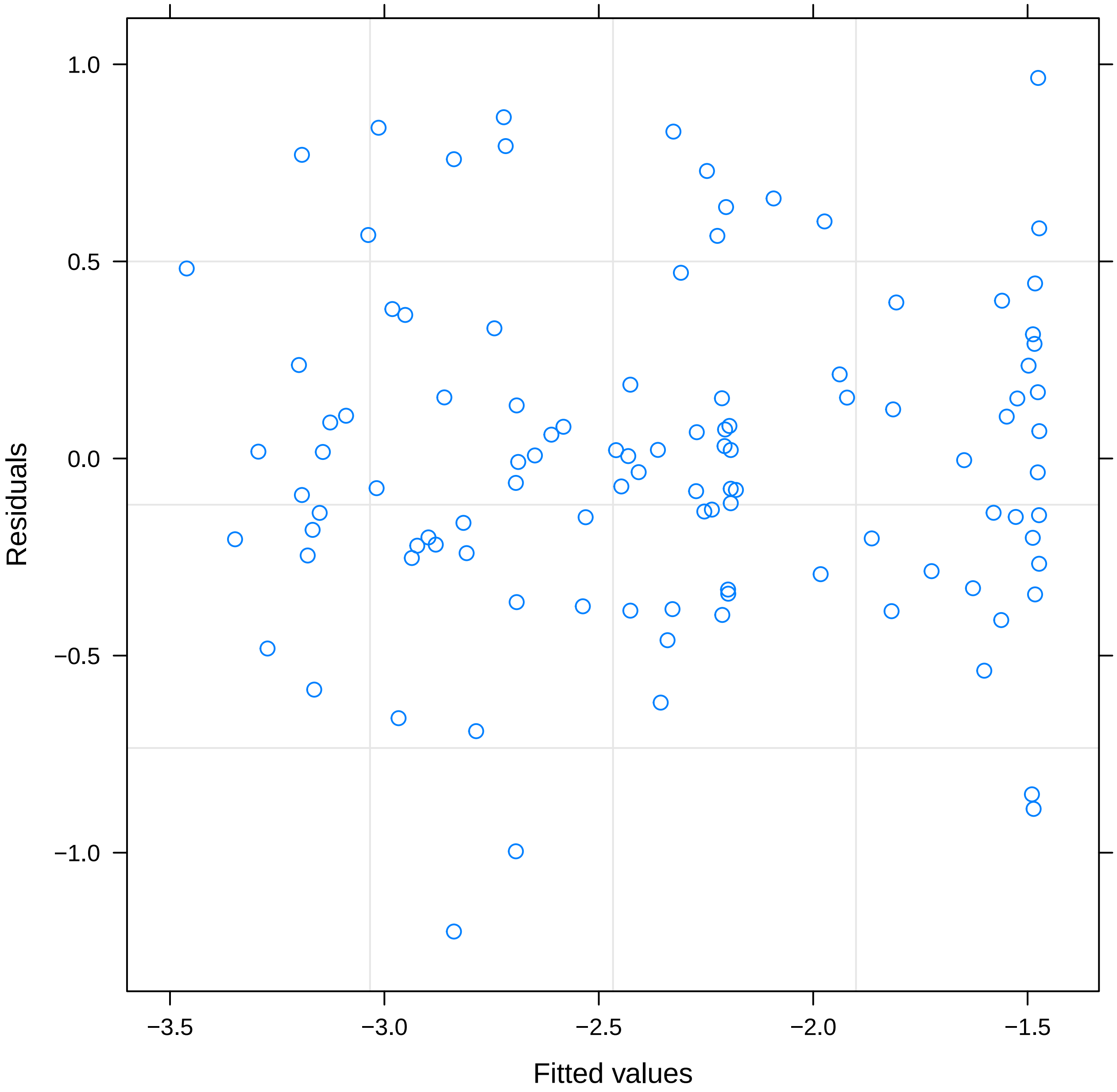


Figure 4: Residuals versus fitted values show no particular pattern that would be concerning.

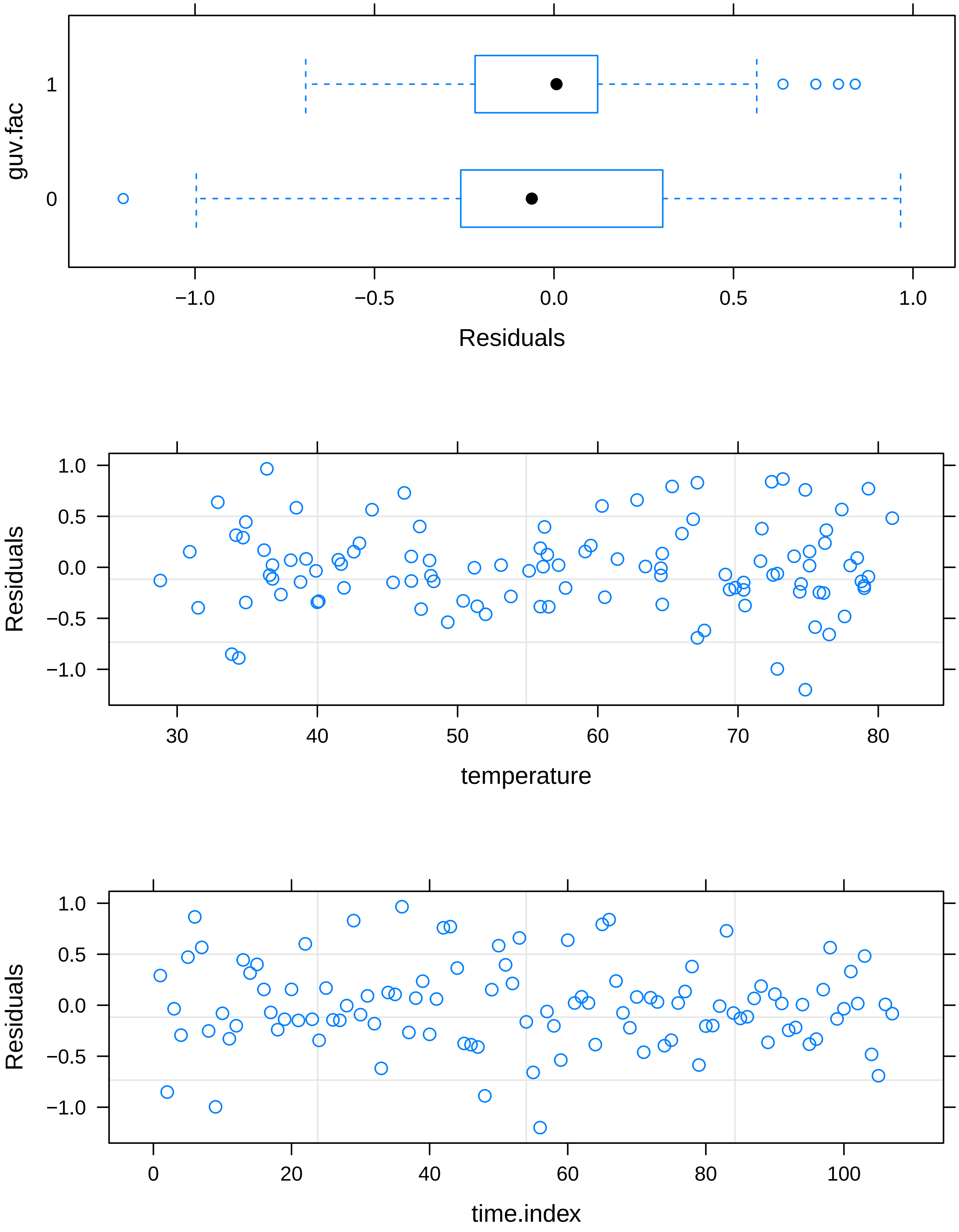


Figure 5: Top: There is slight heteroskedasticity in residuals between pre- and post-GUV periods. Middle: residuals by monthly average temperature show no particular pattern. Bottom: Time participates in the working model only in the serial autocorrelation structure. Residuals by time show no particular pattern.

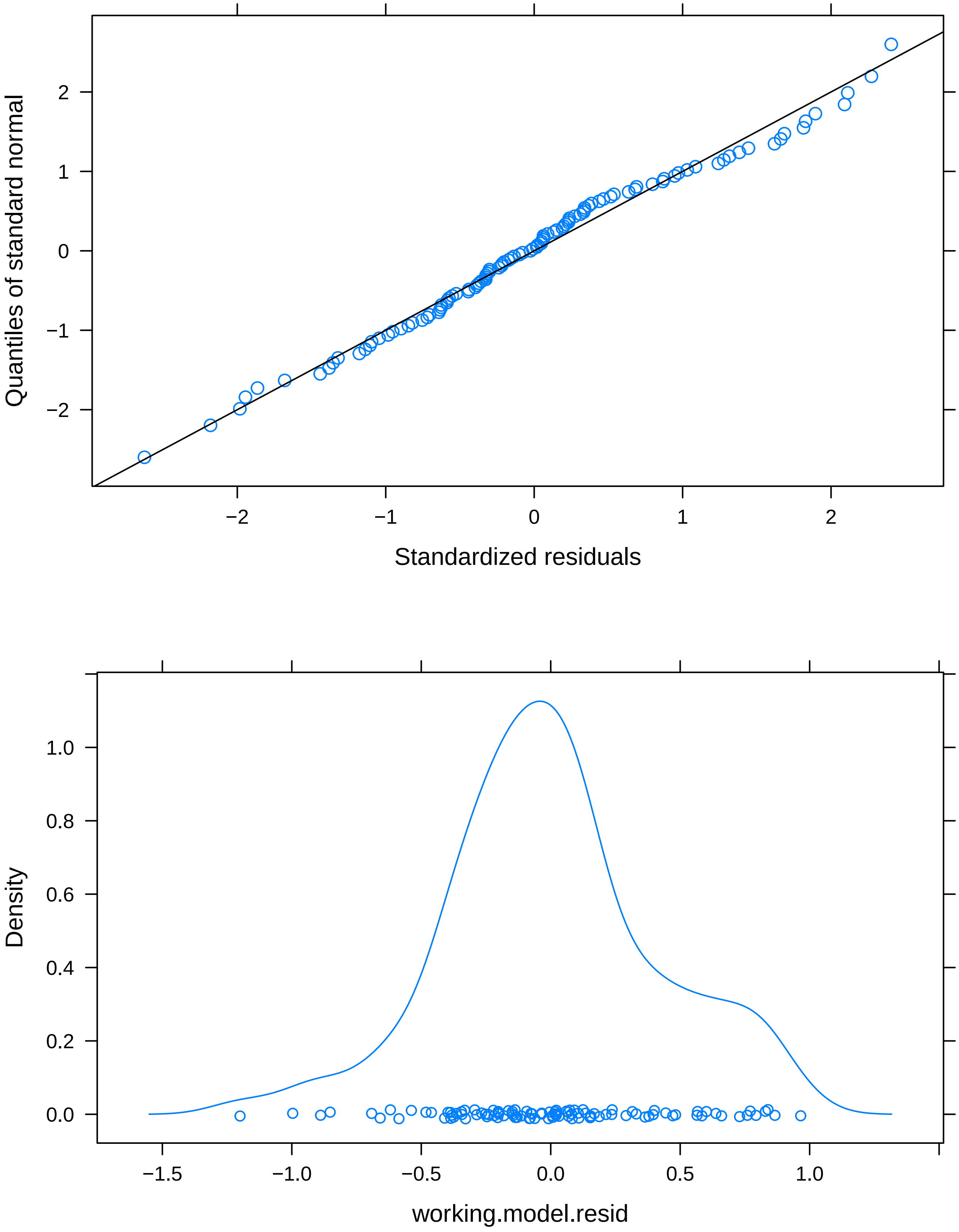


Figure 6: Residuals are a reasonable fit to a Gaussian distribution. Top: qqnormal plot. Bottom: kernel density plot.

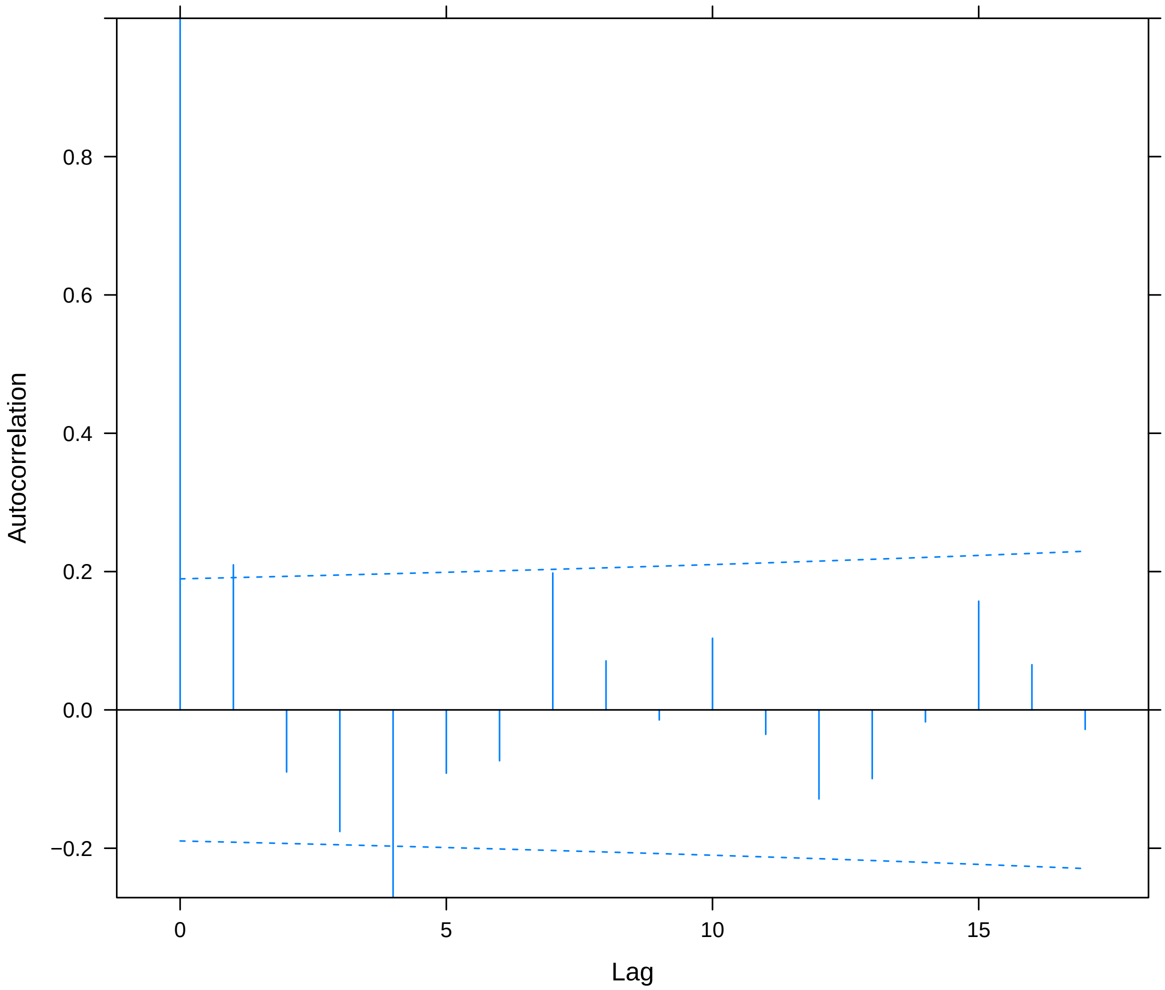


Figure 7: Some residual autocorrelation remains, especially at a lag of four months.
